# Practical Indicators for Risk of Airborne Transmission in Shared Indoor Environments and their Application to COVID-19 Outbreaks

**DOI:** 10.1101/2021.04.21.21255898

**Authors:** Z. Peng, A.L. Pineda Rojas, E. Kropff, W. Bahnfleth, G. Buonanno, S.J. Dancer, J. Kurnitski, Y. Li, M.G.L.C. Loomans, L.C. Marr, L. Morawska, W. Nazaroff, C. Noakes, X. Querol, C. Sekhar, R. Tellier, T. Greenhalgh, L. Bourouiba, A. Boerstra, J.W. Tang, S.L. Miller, J.L. Jimenez

**Affiliations:** Dept. of Chemistry and CIRES; University of Colorado, Boulder, CO, USA; CIMA, UMI-IFAECI/CNRS, FCEyN, Universidad de Buenos Aires—UBA/CONICET, Buenos Aires, Argentina; Leloir Institute—IIBBA/CONICET, CBA, Buenos Aires, Argentina; Dept. of Architectural Engineering, The Pennsylvania State University, University Park, PA, USA; Dept. of Civil and Mechanical Engineering, University of Cassino and Southern Lazio, Italy; Dept. of Microbiology, NHS Lanarkshire, Scotland, UK; School of Applied Sciences, Edinburgh Napier University, Scotland, UK; REHVA Technology and Research Committee, Tallinn University of Technology, Tallinn, Estonia; Dept. of Mechanical Engineering, the University of Hong Kong, Hong Kong, China; Dept. of the Built Environment, Eindhoven University of Technology, Eindhoven, the Netherlands; Dept. of Civil and Environmental Engineering, Virginia Tech, Blacksburg, VA, USA; International Laboratory for Air Quality and Health, Queensland University of Technology, Brisbane, Australia; Dept. of Civil and Environmental Engineering, University of California, Berkeley, CA, USA; School of Civil Engineering, University of Leeds, Leeds, UK; Institute of Environmental Assessment and Water Research, IDAEA, Spanish Research Council, CSIC, Barcelona, Spain; Dept. of the Built Environment, National University of Singapore, Singapore; McGill University, Montreal, QC, Canada; Dept. of Primary Care Health Sciences, University of Oxford, Oxford, UK; The Fluid Dynamics of Disease Transmission Laboratory, Massachusetts Institute of Technology, Cambridge, MA, USA; REHVA (Federation of European Heating, Ventilation and Air Conditioning Associations), BBA Binnenmilieu, the Netherlands; Respiratory Sciences, University of Leicester, Leicester, UK; Dept. Mechanical Engineering, University of Colorado, Boulder, CO, USA

**Keywords:** COVID-19, airborne transmission, indoor air, risk assessment, mitigation

## Abstract

Some infectious diseases, including COVID-19, can be transmitted via aerosols that are emitted by an infectious person and inhaled by susceptible individuals. Most airborne transmission occurs at close proximity and is effectively reduced by physical distancing, but as time indoors increases, infections occur in those sharing room air despite maintaining distancing. There have been calls for quantified models to estimate the absolute and relative contribution of these different factors to infection risk. We propose two indicators of infection risk for this situation, i.e., relative risk parameter (H_r_) and risk parameter (H). They combine the key factors that control airborne disease transmission indoors: virus-containing aerosol generation rate, breathing flow rate, masking and its quality, ventilation and particulate air cleaning rates, number of occupants, and duration of exposure. COVID-19 outbreaks show a clear trend in relation to these factors that is consistent with airborne infection The observed trends of outbreak size (attack rate) vs. H (H_r_) allow us to recommend values of these parameters to minimize COVID-19 indoor infection risk. Transmission in typical pre-pandemic indoor spaces is highly sensitive to mitigation efforts. Previous outbreaks of measles, flu, and tuberculosis were assessed along with recently reported COVID-19 outbreaks. Measles outbreaks occur at much lower risk parameter values than COVID-19, while tuberculosis outbreaks are observed at much higher risk parameter values. Since both diseases are accepted as airborne, the fact that COVID-19 is less contagious than measles does not rule out airborne transmission. It is important that future outbreak reports include information on the nature and type of masking, ventilation and particulate-air cleaning rates, number of occupants, and duration of exposure, to allow us to understand the circumstances conducive to airborne transmission of different diseases.

**Synopsis:** We propose two infection risk indicators for indoor spaces and apply them to COVID-19 outbreaks analysis and mitigation.

## Introduction

Some respiratory infections can be transmitted through the airborne pathway, in which aerosol particles (< 100 μm) are shed by infected individuals and inhaled by others, causing disease in susceptible individuals.^1–4^ It is widely accepted that measles, tuberculosis, and chickenpox are transmitted in this way,^5,6^ and acceptance is growing that this is a major and potentially the dominant transmission mode of COVID-19.^7–13^ There is substantial evidence that smallpox,^14^ influenza,^3^ SARS,^15^ MERS,^6^ and rhinovirus^16^ are also transmitted via aerosols.

There are three airborne transmission scenarios of interest in which infectious and susceptible people: (a) are in close proximity to each other (< 1-2 m), so-called “short-range airborne transmission,”^17^ which is effectively mitigated by physical distancing; (b) share air in the same room, “shared space airborne”; and (c) “longer-distance airborne transmission” in buildings when individuals are not sharing a room (or are in very large rooms), or even between buildings as in the Amoy Gardens SARS outbreak.^15^ Often (b) and (c) are lumped together under “long-range transmission,” but in Scenario (c) transport of pathogen-containing air is more complex, so that the approximation of well mixed air is less valid. It is thus useful to separate these scenarios given the substantial differences in the risk of these situations and actions needed to abate the risk of transmission.

Airborne diseases vary widely in transmissibility, but all of them are most easily transmitted at short range due to the higher concentration of pathogen-containing aerosols close to the infected person. For SARS-CoV-2, a pathogen of initially moderate infectivity (more recently increased by some variants such as Delta), many instances have been reported implicating transmission in shared indoor spaces. Indeed, multiple outbreaks of COVID-19 have been reported in crowded spaces that were relatively poorly ventilated and that were shared by many people for periods of half an hour or longer. Examples include choir rehearsals,^11^ religious services,^18^ buses,^19^ workshop rooms,^19^ restaurants,^4,20^ and gyms,^21^ among others. There are only a few documented cases of longer-distance transmission of SARS-CoV-2, in buildings.^22–24^ However, cases of longer-distance transmission are harder to detect as they require contact tracing teams to have sufficient data to connect cases together and rule out infection acquired elsewhere. Historically, it was only possible to prove longer-distance transmission in the complete absence of community transmission (e.g. ref ^14^).

Being able to quickly assess the risk of infection for a wide variety of indoor environments is of the utmost importance given the impact of the continuing pandemic (and the risk of future pandemics) on so many aspects of life in almost every country of the world. We urgently need to improve the safety of the air that we breathe across a range of environments including child-care facilities, kindergartens, schools, colleges, shops, offices, homes, eldercare facilities, factories, public and private transportation, restaurants, gyms, libraries, cinemas, concert halls, places of worship and mass outdoor events across different climates and socio-economic conditions. There is very little evidence on the actual ventilation rates and the effectiveness of ventilation systems at reducing risks from viral exposure and other indoor pollutants within the majority of buildings. However, data from COVID-19 outbreaks consistently show that a large fraction of buildings worldwide have very low ventilation rates despite the requirements set in national building standards. A host of policy questions – from how to safely re-open schools to how to prevent transmission in high-risk occupational settings – require accurate quantification of the multiple interacting variables that influence airborne infection risk.

Qualitative guidance to reduce the risk of airborne transmission has been published.^25–27^ Different mathematical models have been proposed to help manage risk of airborne transmission,^28,29^ and several models have been adapted to COVID-19.^30–33^ It is important to define quantitative infection risk criteria for different spaces and types of events to more effectively manage the pandemic.^34^ Such criteria could then be used by authorities and policy makers to assist in deciding which activities are permitted under what conditions, so as to limit infection risk across a society. To our knowledge, no such quantitative criteria have been proposed. Besides, often recommendations are complex and vague, e.g., “reduce duration and density of occupancy, and increase ventilation.” However, it is not clear how to combine the different measures together (e.g., is half the duration equivalent to doubling the ventilation?) and it is also not clear what level of mitigation is sufficient to reduce outbreak probability to a low level.

Here, we use a box model to estimate the viral aerosol concentration indoors, and combine it with the Wells-Riley infection model.^28^ The combined model is used to derive two quantitative risk parameters that allow comparing the relative risk of transmission in different situations when sharing room air. We explore the trends in infections observed in outbreaks of COVID-19 and other diseases as a function of these parameters. Finally, we use the parameters to quantify a graphical display of the relative risk of different situations and mitigation options.

## Materials and Methods

### Box model of infection

The box model considers a single enclosed space, in which virus-containing aerosols are assumed to be rapidly uniformly mixed compared with the time spent by the occupants in the space. This is approximately applicable in many situations, but there are some exceptions such as outbreaks where clear directional flow occurred in a room.^18,20^ The mathematical notation used in the paper is summarized in Table S1. The mass balance equation is first written in terms of c, the concentration of infectious quanta in the air in the enclosed space (units of quanta m^-3^). Compared with a model written in terms of aerosol or viral particle concentrations, c has the advantage of implicitly including effects such as the deposition efficiency of the aerosol particles in the lungs of a susceptible person, as well as the efficiency with which such deposited particles may cause infection, the multiplicity of infection, etc. The balance of quanta in the space can be written as:

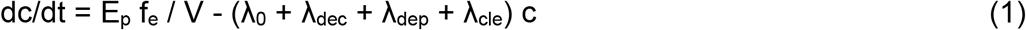

where E_p_ is the emission rate of quanta into the indoor air from an infected person present in the space (quanta / h); f_e_ is the penetration efficiency of virus-carrying particles through masks or face coverings for exhalation (which takes into account the impact of whether the infector wears a face covering); V is the volume of the space; λ_0_ is the first-order rate of removal of quanta by ventilation with outdoor air (h^-1^); λ_cle_ is the removal of quanta by air cleaning devices (e.g. recirculated air with filtering, germicidal UV, portable air cleaners, etc.); λ_dec_ is the infectivity decay rate of the virus; λ_dep_ is the deposition rate of airborne virus-containing particles onto surfaces. E_p_ is a critical parameter that depends strongly on the disease, and it can be estimated with a forward model based on aerosol emission rates and pathogen concentration in saliva and respiratory fluid^31,35^ or by fitting a model such as the one used here to real transmission events.^11,31^

This equation can be solved analytically or numerically for specific situations. Given the enormous number of possible situations, and given the prevalence of outbreaks resulting from longer events where air is shared for a substantial period of time, we consider a sufficiently long event so that a steady state condition (dc/dt ∼ 0) is a reasonable approximation. Writing λ = λ_0_ + λ_cle_ + λ_dec_ + λ_dep_ for simplicity leads to a steady-state infectious quanta concentration of:

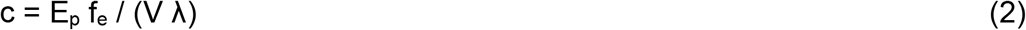

Under the assumption of no infectious quanta at the beginning of the event, a multiplicative factor, r_ss_, can be applied for events too short to approximately reach steady state (see Section S1 for detail) to correct the deviation of the quanta concentration averaged over the event (c_avg_) from that at steady state:

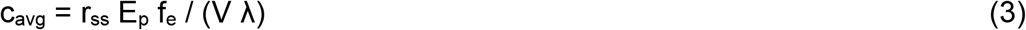

Since the goal is to analyze outbreaks, we assume that only a single infectious person is present in the space, which is thought to be applicable to the outbreaks analyzed below. This allows calculation of the probability of infection, *conditional* to one infectious person being present. The model can also be formulated to calculate the *absolute* probability of infection, if we assume that the probability of an infectious person being present reflects the prevalence of a disease at a given location and time (e.g. refs ^30,36^).

The dose expressed in infectious quanta (n) inhaled by each of the susceptible persons present in the space (N_sus_) is then:

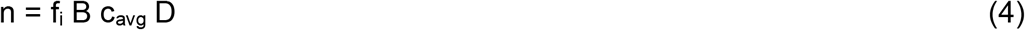

where f_i_ is the penetration efficiency of virus-carrying particles through masks or face coverings for inhalation (which takes into account the effect of the fraction of occupants wearing face coverings); B is the breathing volumetric flow rate of susceptible persons; D is the duration of exposure, assumed to be the same for all the susceptible persons. Substituting:

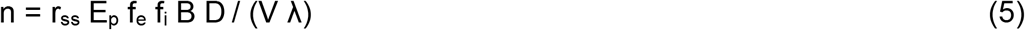

The number of expected secondary infections increases monotonically with increasing n. For an individual susceptible person, by definition of an infectious quantum, the probability of infection is ^28^:

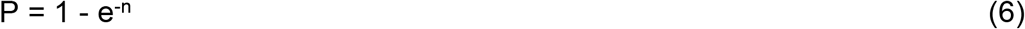

For low values of n, the use of the Taylor expansion for an exponential allows approximating P as:

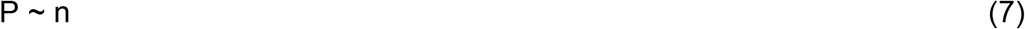

The total number of secondary infections expected, which may also be regarded as the effective reproduction number (R_e_) in a given situation, is then:

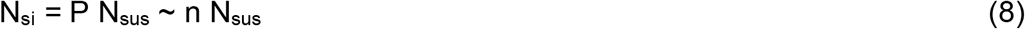

where N_sus_ is the number of susceptible individuals present, which may be lower than the number of occupants due to vaccination or immunity due to past infection. All the outbreaks studied here occurred before COVID-19 vaccines became available. Thus, the number of secondary infections increases linearly with n at lower values, and non-linearly at higher values. We retain the simplified form to define and calculate the risk parameters, but use equation (6) for fitting the outbreak results in Figure 1b below.

**Figure 1:**
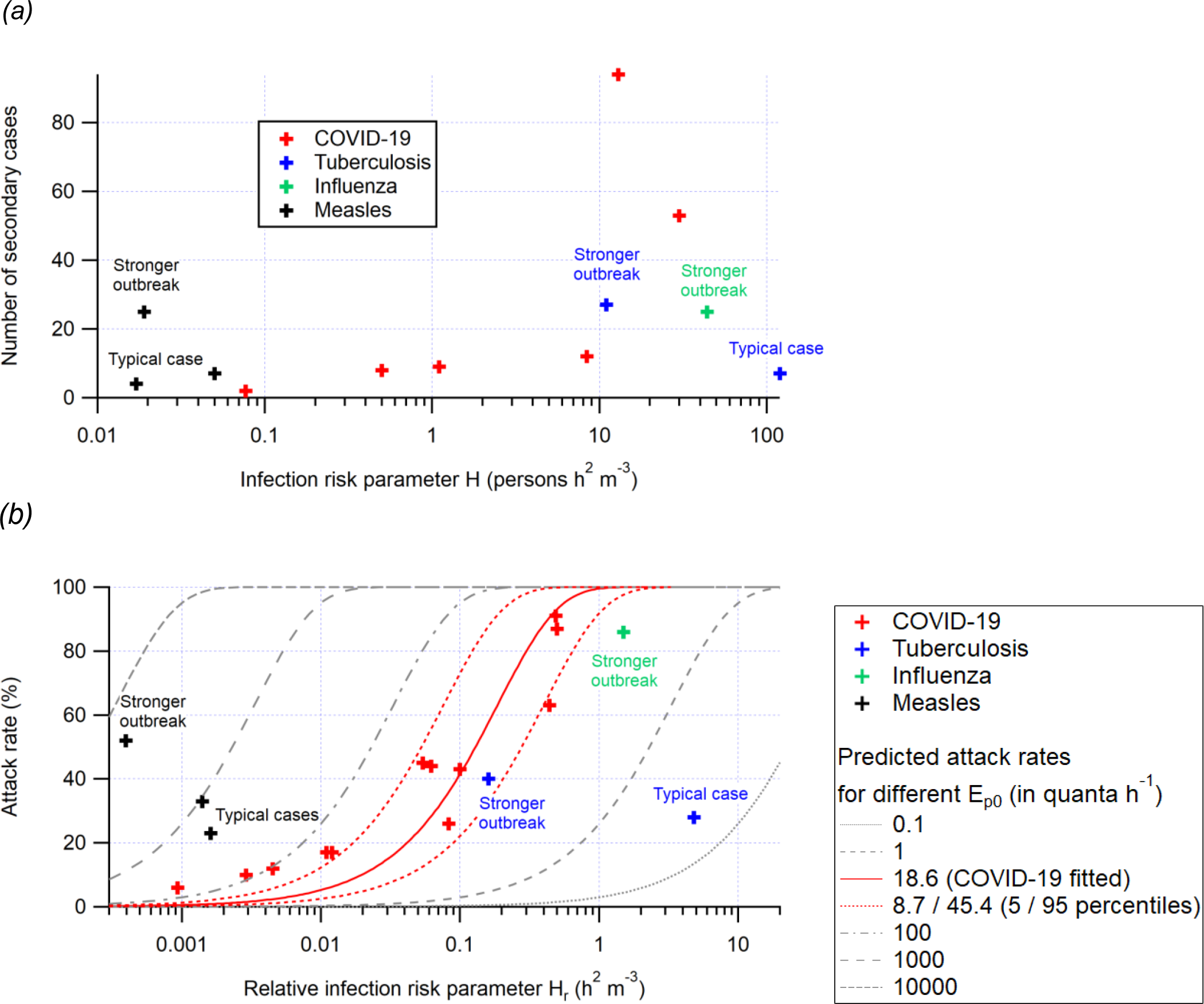
(a) Number of secondary cases vs. the risk parameter H and (b) attack rate vs. the relative risk parameter H_r_ for outbreaks of COVID-19, tuberculosis, influenza, and measles reported in the literature. The fitted trend line of attack rate as a function of H_r_ and its estimated uncertainty range (5^th^ and 95^th^ percentiles) are also shown in (b). All of the outbreaks investigated here involve the original variants of the virus. A variant twice as contagious should result in moving the results to the left by a factor of two or so.

### Risk parameters for airborne infection

We define the relative risk parameter, H_r_, and the risk parameter, H, for airborne infection in a shared space. The purpose of H_r_ and H is to capture the dependency of P and N_si_, respectively, on the parameters that define an event in a given space, in particular those parameters that can be controlled to reduce risk of shared-room airborne transmission. To better capture the controllable actions, E_p_ and B were split into factors that can and cannot be controlled. E_p_ can be expressed as the product E_p0_ x r_E_ where E_p0_ and r_E_ are, respectively, the quanta shedding rate of an infectious person resting and only orally breathing (no vocalization); and the shedding rate enhancement factor relative to E_p0_ for an activity with a certain degree of vocalization and physical intensity (see Table S2a for detail). B can be expressed as B_0_ x r_B_, where B_0_ and r_B_ are the volumetric breathing rate of a sedentary susceptible person in the age group of 41-<51 years (numerically also the average for all age groups) and the relative breathing rate enhancement factor (vs. B_0_) for an activity with a certain physical intensity and for a certain age group (see Table S2b for detail). E_p0_ is uncertain, likely highly variable across the population, and variable over time during the period of infectiousness.^31,35,37^ It may also increase due to new virus variants such as the SARS-CoV-2 Delta variant, that is more contagious, assuming that the increased contagiousness is due to increased viral emission or reduced infectious dose (both of which would increase the quanta emission rate).^38,39^ We note that some variants could in principle also increase transmissibility by lengthening the period of infectiousness for a given person, which by itself would not increase the quanta emission rate in a given situation. B_0_ is relatively well known, and varies with a susceptible person’s age, sex, and body weight, in addition to physical activity level. r_E_ and r_B_ are less uncertain than E_p0_ and are functions of the specific physical and vocalization activities.^31,35,40^ Thus, they are useful in capturing the quantitative impact of specific controllable factors. There could be factors beyond those considered here that lead to variation of viral emissions, such as the respiratory effort of patients with breathing disorders, such as emphysema or asthma.^41^ Such factors can be incorporated into updated Tables in the future.

Then P can be expressed as a function of E_p0_, B_0_ and the product of the other controllable factors as:

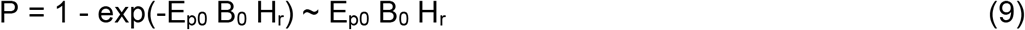

Where H_r_ is the relative risk parameter:

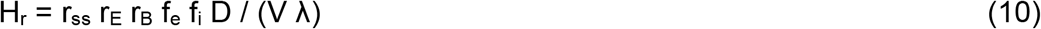

It has a unit of h^2^ m^-3^, which indicates the increase of risk with duration (h), the inverse of the ventilation rate expressed as air changes per hour (1/h^-1^ = h), and the inverse of the volume of the space (1/m^3^). The other parameters such as the effect of masking are dimensionless. H_r_ includes the relative increase of the emission with activity (r_E_), but not the quanta emission rate by a resting and orally breathing infector (E_p0_). This allows using the same risk parameters for different diseases, which will naturally separate in the graphs according to their transmissibility. The four terms that make up λ may vary in relative importance for different diseases and conditions. λ_dec_ ∼ 1.1 h^-1 42^ has been reported for COVID-19. λ_dec_ depends on temperature and relative humidity.^43,44^ λ_dep_ depends on particle size and the geometry and airflow in a given space. Respiratory particle sizes in the range from 1-5 μm are thought to play a role in aerosol transmission of COVID-19, due to a combination of high emission rates by activities such as talking^45^ and low deposition rates. λ_dep_ for a typical furnished indoor space span 0.2-2 h^-1^ over this size range, with faster deposition for larger particles.^46^ λ_0_ varies from ∼12 h^-1^ for airborne infection isolation rooms,^47^ ∼6 h^-1^ for laboratories, ∼0.5 h^-1^ for residences,^48^ and ∼1 h^-1^ for offices.^1,49^ Very little ventilation data is available for many semi-public spaces such as shops, restaurants and bars or transportation. λ_cle_ can vary from 0, if such systems are not in use, to several h^-1^ for adequately sized systems. Ventilation with clean outdoor air will be important in most situations, while virus decay and deposition likely contribute but are more uncertain for COVID-19, based on current information. In particular, the size distribution of aerosols containing infectious viruses is uncertain.

We consider a worst-case scenario where rates of deposition and infectivity decay are small compared with ventilation and air cleaning and can therefore be neglected. This also allows using the same relative risk parameter to compare different airborne diseases. This yields:

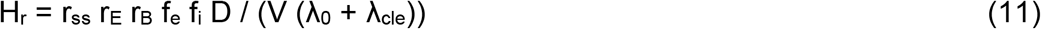

H_r_ can be recast as:

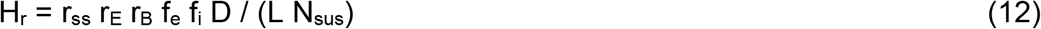

where:

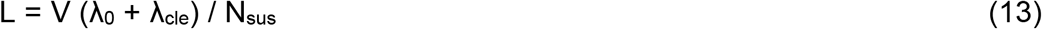

Assuming that all of the people present in a space are susceptible to infection, L is equivalent to the ventilation plus air cleaning rate per person present in the space, (typically expressed in liters s^-1^ person^-1^ in standards and guidelines such as from refs ^50–52^. If some fraction of the people present are immune to the disease, then L is larger than the corresponding personal ventilation rate in the guidance. While this recasting will be useful to persons familiar with ventilation guidelines, we keep the form in equation (11) for most further analyses, since the number of people allowed in a space is one of the critical variables that can be examined with this relative risk parameter.

We insert equation (9) into equation (8) and obtain

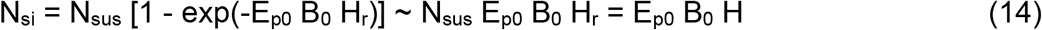

where we define the risk parameter

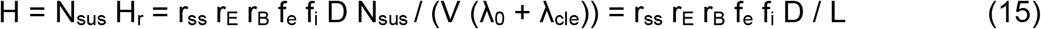

When there is only one infector present and n is small, H is proportional to N_si_, i.e., the outbreak size. When it is unknown whether an infector is present, H is an approximate indicator of the absolute probability of infection (P_a_), since the expected value of number of infectors (N_i_) is the product of number of occupants (N) and probability of an occupant being infectious, a measure of prevalence of infectious people in local population (η_I_), as shown below:

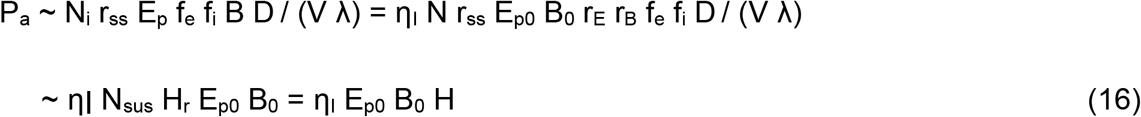

The precise estimation of η_I_ is complex for several reasons. Most transmission may be associated with those infected with high viral loads,^53,54^ as many infected people may not shed virus into the air.^37^ Also much spread is by people with few or no symptoms who may not know that they are infected,^55^ however a fraction of symptomatic infectious people are typically in isolation. There is also significant variation in viral load and infectivity during the course of the disease.^56^ For these reasons, it is very difficult to determine η_I_ precisely based on test data. For a situation with multiple potential infectors present (e.g., a COVID ward in a hospital), the risk parameters should be multiplied by the number of infectors.

For the analysis later in the paper that does not involve activity type or face covering choice (and thus does not involve r_E_, r_B_, f_e_, or f_i_), we define another parameter (H’), which is closely related to H:

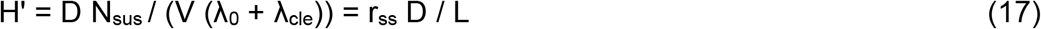

This parameter only captures the characteristics of susceptible people’s presence in the indoor space, not of their behavior.

### Value of the risk parameters for documented outbreaks of COVID-19

An important advantage of the simple risk parameters is that their values can be calculated for outbreaks that are documented in the scientific literature. Values for documented COVID-19 outbreaks are shown in Table 1 (r_E_ and r_B_ are estimated based on the likely types of activities in each case,^31,35,40^ see Table S2 for typical values). Also included are values for outbreaks documented in the literature for tuberculosis and measles, which are widely accepted to transmit through the air, and an influenza outbreak that was clearly due to airborne transmission. We have included all the outbreaks known to us for which sufficient information was available to estimate airborne infection risk. Most public health investigations so far in this pandemic have neglected to report ventilation rates, the volume of the space, filter and air cleaner efficiencies, and other building science details, and thus their airborne risk cannot be estimated. It is important that future outbreak reports include this information, to allow expanding our knowledge of the circumstances conducive to airborne transmission of different diseases.

**Table 1:**
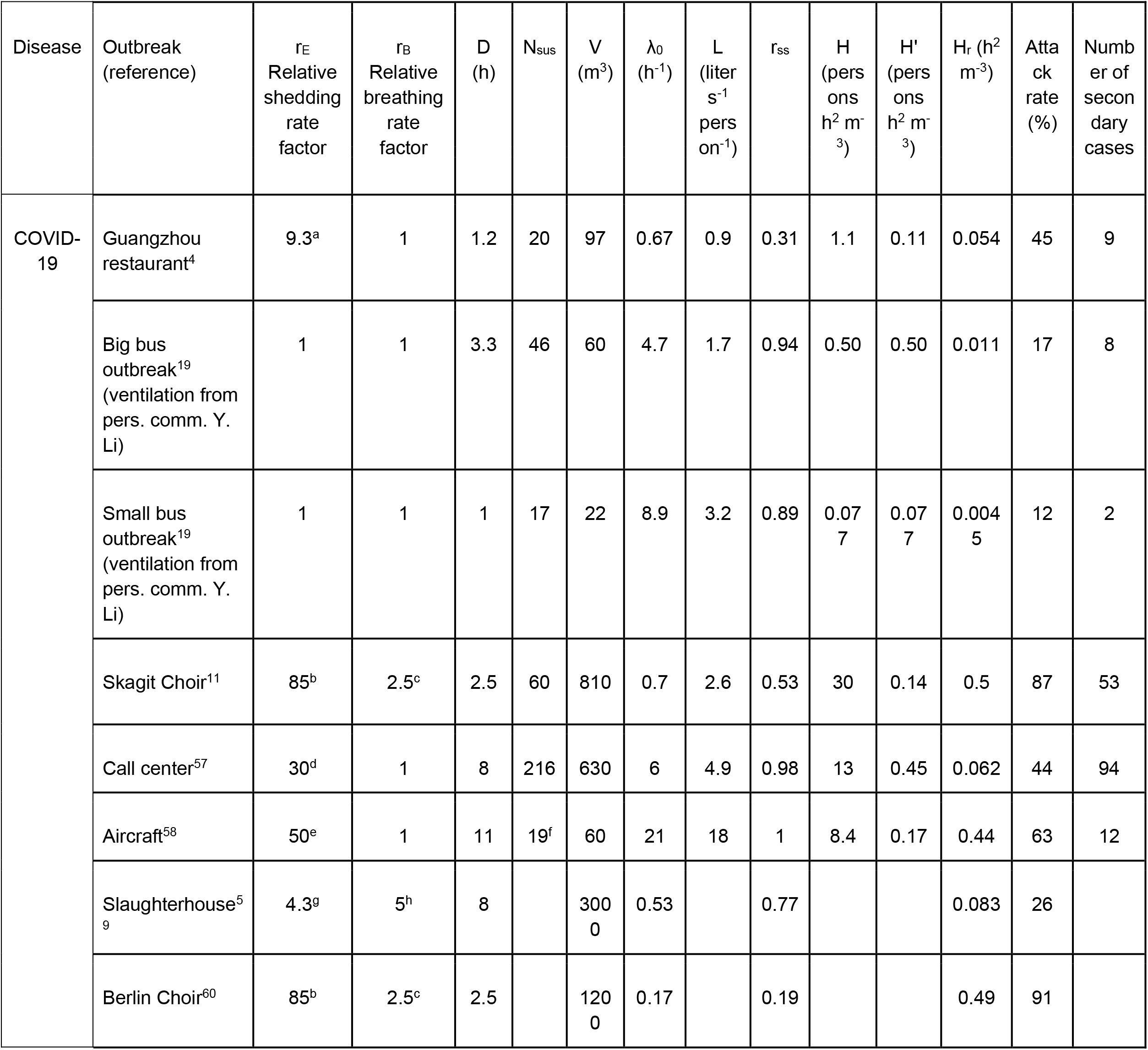
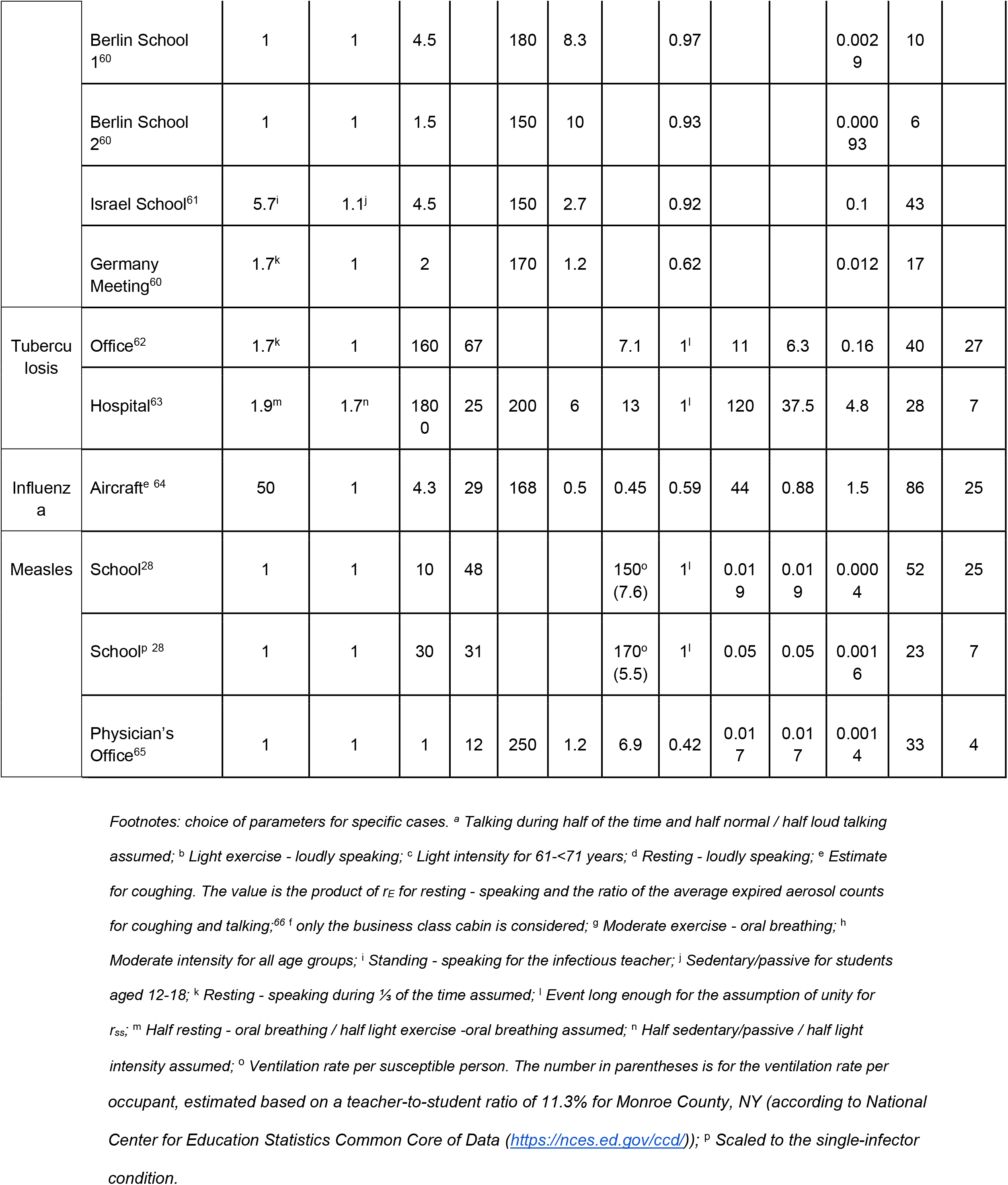
Parameters for outbreaks documented in the literature. Missing parameters could not be calculated from the information given in the literature references.

We see that the COVID-19 outbreaks that have been documented span ∼2.5 orders of magnitude range of the risk parameter H ∼ 0.09-30 persons h^2^ m^-3^. Numbers of secondary cases of these outbreaks generally increase with H (Figure 1a). Aiming to maintain values far below the threshold of 0.05 persons h^2^ m^-3^ should help reduce outbreaks.

H_r_ correlates well with the attack rates for the outbreaks reported in Table 1 (Figure 1b, H_r_ can be calculated for more literature outbreaks than H). COVID-19 outbreaks are observed for H_r_ > 0.001 h^2^ m^-3^, and thus indoor activities should be limited to conditions below this value during the pandemic whenever possible.

A trend line can be fitted to the attack rate vs. H_r_ dataset with the Box/Wells-Riley model (Figure 1b), with the fitting parameter being E_p0_, i.e. the basic quanta shedding rate (when breathing only, no vocalization). A best-fit E_p0_ of 18.6 quanta h-1 was obtained (with B_0_ = 0.288 m^3^ h^-1^ assumed for all occupants for simplicity). When the uncertainties of H_r_ and attack rates are considered, the uncertainty of E_p0_ can also be estimated through Monte Carlo uncertainty propagation, described in detail in Section S2. The 5^th^ and 95^th^ percentiles of the E_p0_ values are 8.7 and 45.4 quanta h^-1^, respectively. This range is higher than that suggested by Buonanno et al. (2 quanta h^-1^),^31,35^ but overlapping with the uncertainty range provided by those authors. Note that outbreaks are typically only observed for individuals with higher quanta emission rates; many infected individuals have low emission rates and the risk in those situations will be lower than estimated here.^31,35^ The attack rates estimated according to this trend line have a high correlation with the actual attack rates (r^2^ = 0.90; Figure S1). Given the small size of the dataset of H_r_ and attack rates, we cross-validate this fitting by the leave-one-out method.^67^ The 12 values of E_p0_ obtained in the cross validation have maximum and mean relative absolute deviations of 20% and 6%, respectively, from the best-fit value, showing the robustness of the fitting.

Close alignment supports the dominant shared-room airborne character of these COVID-19 outbreaks. If the outbreaks had major components of other transmission modes (e.g., fomite or close-range transmission), we would expect a dependence on other parameters not considered here and overall much lower correlation with the risk parameters. Nevertheless, it can be observed that the attack rates of the outbreaks at low H_r_ (∼0.01 h^2^ m^-3^ or lower) are higher than the fitted curve. This can be due to several factors, including i) other transmission routes (e.g., short-range airborne transmission, for which the risk cannot be well captured by the model in this study and can still be significant at low H_r_) or ii) the detection of only outlier cases of long-range airborne transmission resulting from the variability of E_p0_ (e.g., cases where the infector’s actual quanta emission rate was extremely high), as the other cases may have too few secondary infections to be documented in the literature.

### Effect of building parameters vs. human activities

The type of activity performed in each case (captured by the product of r_E_ and r_B_) contributes substantially to the difference in H between these cases. When human activities are not taken into account, the parameter H’ only spans a narrow range of 0.09-0.56 person h^2^ m^-3^. This is probably due to similar per-person ventilation rates in many public indoor spaces (on the order of a few liter s^-1^ person^-1^)^50^ and similar lengths of common events (in hours).

Similar to H, the variation in the values of H_r_ for the outbreaks in Table 1 is also largely determined by r_E_ and r_B_. If they are not taken into account, H_r_ for all outbreaks would vary in the narrow range 0.001 to ∼0.01 h^2^ m^-3^, as V and λ_0_ are building characteristics and D, as discussed above, is usually in hours. This implies that, in the presence of a single infector, reducing vocalization and/or physical intensity levels of the indoor activity is a very effective way to lower the infection risk of susceptible individuals. Reducing event length can also help, while reducing occupancy cannot in this case, as shown by equation (10).

It is possible that some of the most visible outbreaks are associated with super-emitter individuals, who shed virus particles at higher rates than others.^41,68–70^ If that is the case, the actual H_r_ values at which significant transmission starts to appear in the presence of individuals that are not super emitters may be higher than those determined here. However, if super-emitters are important, so will be their contribution to total spread, and thus one should try to reduce the risk to reduce the probability of such events occurring.

### Values of the risk parameters for outbreaks of other airborne diseases

In Table 1 and Figure 1 we also include a few reported indoor outbreaks of three other diseases with significant airborne transmission, i.e., tuberculosis, influenza, and measles. For outbreaks to have a similar number of secondary cases or attack rate, H or H_r_ needs to be higher for tuberculosis and influenza and lower for measles than that for COVID-19 (Figure 1). Note that many of the children present in the measles outbreak were vaccinated, but the risk parameter framework can still be applied by considering the number of susceptible children present. This difference is mainly due to differences in E_p0_ (lower for tuberculosis and influenza and higher for measles; Figure 1b). A higher E_p0_ for measles may indicate a larger amount of airborne measles virus in breath or a steeper dose-response curve for the measles virus than SARS-CoV-2, or both. A novel disease as contagious as measles would make almost any indoor situation prone to superspreading. On the other hand, tuberculosis and influenza are less contagious.

Tuberculosis transmission is propagated because untreated infected people remain contagious for years.^71^ The influenza outbreak occurred in an airplane without ventilation with the index case constantly coughing, and represents an extreme for this disease.^64^ Most influenza patients emit significantly less virus.^72^ More discussions about quanta emission rates of different diseases can be found in refs ^35,73^.

Given that both the measles and tuberculosis pathogens are widely accepted as airborne, the intermediate risk profile for COVID-19 in Figure 1b is not inconsistent with airborne transmission, contrary to frequently made arguments.^74,75^ Contagiousness of a disease does not necessarily indicate the transmission route.^76^ Airborne diseases can vary in their contagiousness depending on parameters such as the amount of virus shed, the survival of the virus in the air, the dose-response relationship for infection, and other parameters. The only fundamental requirement is that transmission needs to be sufficient for the disease to survive as such, something COVID-19 has had no trouble with so far.

### Graphical representation of relative risks of different situations

When it is not known whether infectors are present at an indoor event, all occupants must be considered possible infectors. We assume that the probability of an occupant being infectious is the same as the fraction of infectious people in the local population (η_I_). H indicates the risk of an outbreak. Consequently, the risk also depends on the number of occupants in addition to vocalization level, event duration, ventilation, and mask wearing. Jones et al.^25^ estimated the dependency of the infection risk on these factors and tabulated it in a manner similar to Table 2. However, they only did so qualitatively. Having defined H as a risk parameter, we can assess the risk more quantitatively based on H values (as well as contact times allowed until outbreak risk is significant (H = 0.05 persons h^2^ m^-3^) and attack rates) under different conditions (Table 2). Although the actual risk also depends on η_I_ and the choice of the threshold for high risk (red cells in Table 2) is subjective, the risk parameter (H value) in Table 2 seems to vary in a smaller range than the corresponding table in Jones et al..^25^ We also show that being outdoors (with much better ventilation than indoors) has a greater expected benefit than they estimated.

**Table 2:**
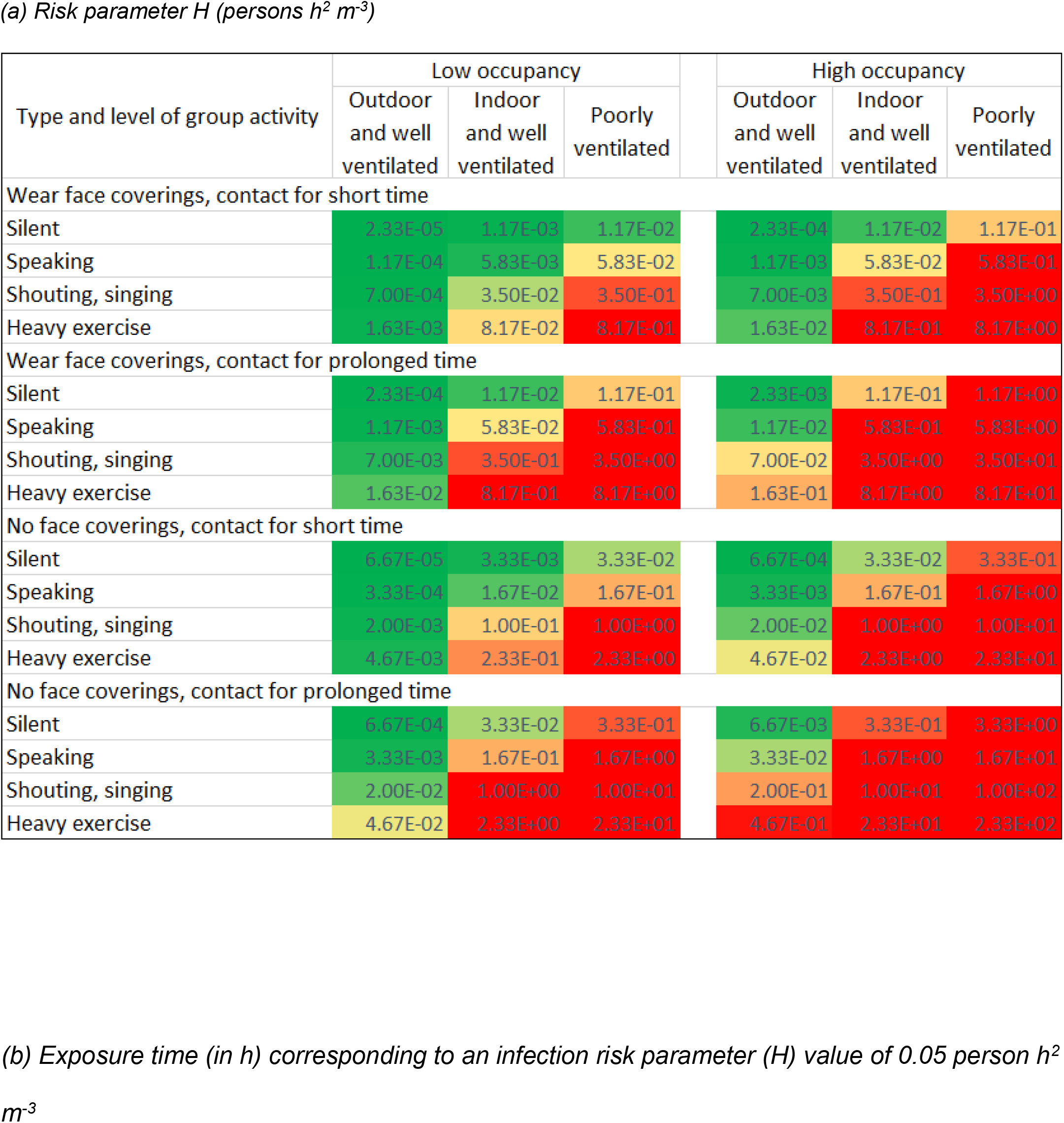
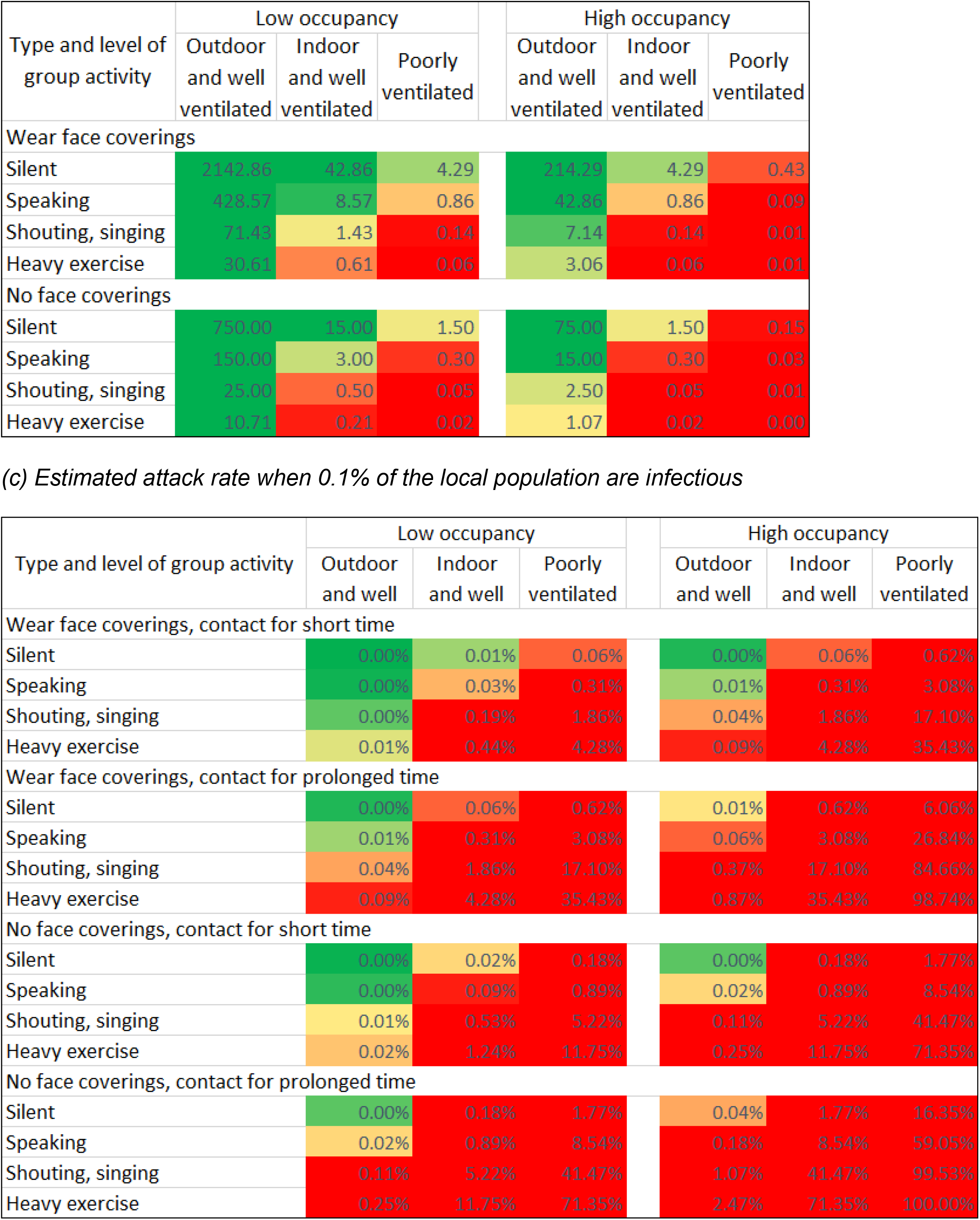
(a) Values of the airborne infection risk parameter (H, in persons h^2^ m^-3^), (b) exposure times corresponding to H = 0.05 persons h^2^ m^-3^, and (c) predicted attack rates with 0.1% infectious people in local population in shared spaces under different conditions in the similar format of Fig. 3 of ref 25. An additional type of activity (“heavy exercise”) is included. Table S3 details the specific choices of the conditions in Table 2. Note that these specifications can be changed as needed, which is easy to implement in the COVID-19 Aerosol Transmission Estimator (Figure S2). Color of a cell varies (a) with H value from green (0 persons h^2^ m^-3^) via yellow (0.05 persons h^2^ m^-3^) to red (≥0.5 persons h^2^ m^-3^), (b) with exposure time from red (0.1 h) via yellow (1 h) to green (10 h), and (c) with predicted attack rate from green (0) via yellow (0.01%) to red (0.1%). The selection of the colors in Table 2a was based on the following considerations: i) no risk (H=0 persons h^2^ m^-3^) for green; ii) no documented outbreaks when H<0.05 persons h^2^ m^-3^ (Figure 1a) (thus 0.05 persons h^2^ m^-3^ for yellow); iii) outbreaks with significant numbers of secondary infections when H≥0.5 persons h^2^ m^-3^ (Figure 1a) (thus red). For that in Table 2b, relatively simple numbers are chosen for the thresholds that correspond to the thresholds for H in Table 2a. As probability of infection is given in Table 2c, its colors are chosen based on the personal risk tolerance of the authors. These tables are available in the online transmission risk estimator, and all of their aspects can be modified depending on specific situations and preferences.

| Type and level of group activity | Low occupancy |  |  |  | High occupancy |  |  |
| --- | --- | --- | --- | --- | --- | --- | --- |
|  | Outdoor and well ventilated | Indoor and well ventilated | Poorly ventilated |  | Outdoor and well ventilated | Indoor and well ventilated | Poorly ventilated |
| Wear face coverings, contact for short time |  |  |  |  |  |  |  |
| Silent | 2.33E-05 | 1.17E-03 | 1.17E-02 |  | 2.33E-04 | 1.17E-02 | 1.17E-01 |
| Speaking | 1.17E-04 | 5.83E-03 | 5.83E-02 |  | 1.17E-03 | 5.83E-02 | 5.83E-01 |
| Shouting, singing | 7.00E-04 | 3.50E-02 | 3.50E-01 |  | 7.00E-03 | 3.50E-01 | 3.50E+00 |
| Heavy exercise | 1.63E-03 | 8.17E-02 | 8.17E-01 |  | 1.63E-02 | 8.17E-01 | 8.17E+00 |
| Wear face coverings, contact for prolonged time |  |  |  |  |  |  |  |
| Silent | 2.33E-04 | 1.17E-02 | 1.17E-01 |  | 2.33E-03 | 1.17E-01 | 1.17E+00 |
| Speaking | 1.17E-03 | 5.83E-02 | 5.83E-01 |  | 1.17E-02 | 5.83E-01 | 5.83E+00 |
| Shouting, singing | 7.00E-03 | 3.50E-01 | 3.50E+00 |  | 7.00E-02 | 3.50E+00 | 3.50E+01 |
| Heavy exercise | 1.63E-02 | 8.17E-01 | 8.17E+00 |  | 1.63E-01 | 8.17E+00 | 8.17E+01 |
| No face coverings, contact for short time |  |  |  |  |  |  |  |
| Silent | 6.67E-05 | 3.33E-03 | 3.33E-02 |  | 6.67E-04 | 3.33E-02 | 3.33E-01 |
| Speaking | 3.33E-04 | 1.67E-02 | 1.67E-01 |  | 3.33E-03 | 1.67E-01 | 1.67E+00 |
| Shouting, singing | 2.00E-03 | 1.00E-01 | 1.00E+00 |  | 2.00E-02 | 1.00E+00 | 1.00E+01 |
| Heavy exercise | 4.67E-03 | 2.33E-01 | 2.33E+00 |  | 4.67E-02 | 2.33E+00 | 2.33E+01 |
| No face coverings, contact for prolonged time |  |  |  |  |  |  |  |
| Silent | 6.67E-04 | 3.33E-02 | 3.33E-01 |  | 6.67E-03 | 3.33E-01 | 3.33E+00 |
| Speaking | 3.33E-03 | 1.67E-01 | 1.67E+00 |  | 3.33E-02 | 1.67E+00 | 1.67E+01 |
| Shouting, singing | 2.00E-02 | 1.00E+00 | 1.00E+01 |  | 2.00E-01 | 1.00E+01 | 1.00E+02 |
| Heavy exercise | 4.67E-02 | 2.33E+00 | 2.33E+01 |  | 4.67E-01 | 2.33E+01 | 2.33E+02 |

(b) Exposure time (in h) corresponding to an infection risk parameter (H) value of 0.05 person h<sup>2</sup> m<sup>-3</sup>
| Type and level of group activity | Low occupancy |  |  | High occupancy |  |  |
| --- | --- | --- | --- | --- | --- | --- |
|  | Outdoor and well ventilated | Indoor and well ventilated | Poorly ventilated | Outdoor and well ventilated | Indoor and well ventilated | Poorly ventilated |
| Wear face coverings |  |  |  |  |  |  |
| Silent | 2142.86 | 42.86 | 4.29 | 214.29 | 4.29 | 0.43 |
| Speaking | 428.57 | 8.57 | 0.86 | 42.86 | 0.86 | 0.09 |
| Shouting, singing | 71.43 | 1.43 | 0.14 | 7.14 | 0.14 | 0.01 |
| Heavy exercise | 30.61 | 0.61 | 0.06 | 3.06 | 0.06 | 0.01 |
| No face coverings |  |  |  |  |  |  |
| Silent | 750.00 | 15.00 | 1.50 | 75.00 | 1.50 | 0.15 |
| Speaking | 150.00 | 3.00 | 0.30 | 15.00 | 0.30 | 0.03 |
| Shouting, singing | 25.00 | 0.50 | 0.05 | 2.50 | 0.05 | 0.01 |
| Heavy exercise | 10.71 | 0.21 | 0.02 | 1.07 | 0.02 | 0.00 |

| Type and level of group activity | Low occupancy |  |  | High occupancy |  |  |
| --- | --- | --- | --- | --- | --- | --- |
|  | Outdoor and well | Indoor and well | Poorly ventilated | Outdoor and well | Indoor and well | Poorly ventilated |
| Wear face coverings, contact for short time |  |  |  |  |  |  |
| Silent | 0.00% | 0.01% | 0.06% | 0.00% | 0.06% | 0.62% |
| Speaking | 0.00% | 0.03% | 0.31% | 0.01% | 0.31% | 3.08% |
| Shouting, singing | 0.00% | 0.19% | 1.86% | 0.04% | 1.86% | 17.10% |
| Heavy exercise | 0.01% | 0.44% | 4.28% | 0.09% | 4.28% | 35.43% |
| Wear face coverings, contact for prolonged time |  |  |  |  |  |  |
| Silent | 0.00% | 0.06% | 0.62% | 0.01% | 0.62% | 6.06% |
| Speaking | 0.01% | 0.31% | 3.08% | 0.06% | 3.08% | 26.84% |
| Shouting, singing | 0.04% | 1.86% | 17.10% | 0.37% | 17.10% | 84.66% |
| Heavy exercise | 0.09% | 4.28% | 35.43% | 0.87% | 35.43% | 98.74% |
| No face coverings, contact for short time |  |  |  |  |  |  |
| Silent | 0.00% | 0.02% | 0.18% | 0.00% | 0.18% | 1.77% |
| Speaking | 0.00% | 0.09% | 0.89% | 0.02% | 0.89% | 8.54% |
| Shouting, singing | 0.01% | 0.53% | 5.22% | 0.11% | 5.22% | 41.47% |
| Heavy exercise | 0.02% | 1.24% | 11.75% | 0.25% | 11.75% | 71.35% |
| No face coverings, contact for prolonged time |  |  |  |  |  |  |
| Silent | 0.00% | 0.18% | 1.77% | 0.04% | 1.77% | 16.35% |
| Speaking | 0.02% | 0.89% | 8.54% | 0.18% | 8.54% | 59.05% |
| Shouting, singing | 0.11% | 5.22% | 41.47% | 1.07% | 41.47% | 99.53% |
| Heavy exercise | 0.25% | 11.75% | 71.35% | 2.47% | 71.35% | 100.00% |

Note that, although occupancy has no impact on attack rate if an infector is present, occupancy affects the risk in two ways when the presence of infector(s) is unknown, i.e., i) the probability of the presence of an infector in a certain locality and ii) the size (number of secondary cases) of the outbreak if it occurs. Therefore, lowering occupancy has double benefits.

### Risk evaluation for indoor spaces with pre-pandemic and mitigation scenarios

Values of the risk parameter H for some typical public spaces under pre-pandemic conditions are tabulated in Table S4 and shown in Figure 2. H in all pre-pandemic settings is on the order of 0.05 persons h^2^ m^-3^ or higher, implying significant risk of outbreak during the pandemic. Often, ventilation rates may be lower than official guidance due to e.g. malfunction, lack of maintenance, or attempts to save energy. Substandard ventilation, coupled with poor air distribution, substantially increases the risk of an outbreak and its size. This is consistent with observations that indicate that COVID-19 outbreaks are disproportionately observed in poorly-ventilated environments, which are specifically spaces with little to no added outdoor air or adequately filtered air.^77^However, H and H_r_ of all of the pre-pandemic spaces are in a regime highly sensitive to mitigation efforts. Therefore mitigation measures such as increasing ventilation or air cleaning, reducing voice volume when speaking, reducing occupancy, shortening duration of occupancy and mask wearing are required to reduce the risk of transmission in similar settings. With mitigation measures implemented, H in these settings can be lowered to the order of 0.01 persons h^2^ m^-3^, low enough to avoid major outbreaks. Particularly, for hospital general examination room, a high-risk setting where there can be coughing infectors emitting quanta at a high rate, a combination of an improvement of ventilation rate to 6 h^-1^, application of higher efficiency air filters, a halved duration, and a requirement of fit-tested N95 respirators can lower the attack rate from ∼90% to negligible (Figure 2c). Use of high-quality masks (e.g., N95/FFP3) is highly effective among the mitigation measures.

**Figure 2:**
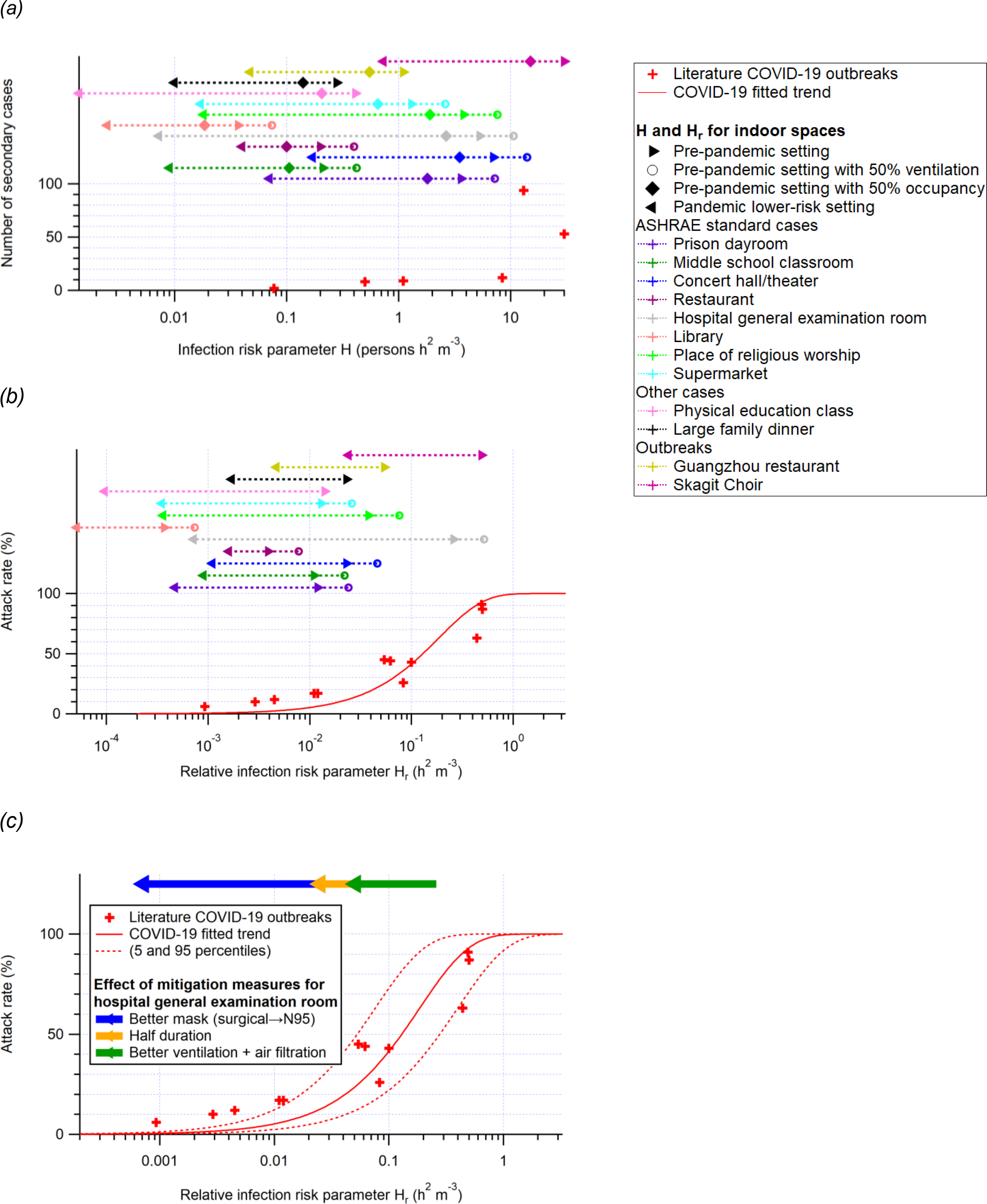
(a, b) same format as Figure 1, but for COVID-19 only. Also shown are the H and H_r_ values for several common indoor situations listed in Table S4. The H values for the cases with pre-pandemic settings except for a lower occupancy and the H and H_r_ values for the ASHRAE standard cases^50^ with pre-pandemic settings except for a lower ventilation rate are shown for comparison. The standalone legend box is for (a) and (b) only. (c) The approximately multiplicative effects of various mitigation measures for the hospital general examination room case are also shown as an example.

### Calculation of risk parameters for specific situations

The calculation of H, H’, and H_r_ for specific situations of interest has been implemented in the COVID-19 aerosol transmission estimator, which is freely available online.^30^ The estimator is a series of spreadsheets that implement the same aerosol transmission model described by Miller et al.^11^ It allows the user to make a copy into an online Google spreadsheet or download it as a Microsoft Excel file for adaptation to the situations of interest to each user. The model can be used to estimate the risk of specific situations, to explore the reduction in transmission due to different control measures (e.g. increased ventilation, masking etc.), and to understand aerosol transmission modeling for incorporation into more complex models. The model also allows the estimation of the average CO_2_ concentration during an activity, as an additional indicator of indoor risk, and to facilitate the investigation of the relationship between infection risk and CO_2_ concentrations.^36,78^ A screenshot of the estimator is shown in Figure S2.

In addition a sheet allows recalculating Table 2 in this paper for sets of parameters different from those used here (and shown in Table S2).

## Discussion

We have explored the relationship between airborne infection transmission when sharing indoor spaces and the parameters of the space using a box / Wells-Riley model. We have derived an expression for the number of secondary infections, and isolated the controllable terms in this expression in two airborne transmission risk parameters, H and H_r_.

We find a consistent relationship, with increasing attack rate in the known COVID-19 outbreaks, as the value of H_r_ increases. This provides some confidence that airborne transmission is important in these outbreaks, and that the models used here capture the key processes important for airborne transmission.

Outbreaks have been observed when H_r_ is on the order of 0.001 h^2^ m^-3^ and higher. A criterion based on H_r_ (e.g. H_r_ > 0.01 or 0.1 h^2^ m^3^, depending on available contact tracing resources) can also be used to determine if an occupant of an indoor space is a “close contact” of an identified infector in the same space through the shared-room air airborne transmission (note that this criterion does not apply to close contacts at risk of infection through short-range (<2 m) airborne transmission). The lowest H for the major COVID-19 outbreaks in indoor settings reported in the literature is ∼0.1 person h^2^ m^-3^. Note that all the outbreaks investigated here concern the early to mid-2020 variants of SARS-CoV-2, and a variant twice as contagious as those should reduce the tolerable values of the parameters by about a factor of 2. H can be orders of magnitude higher for the superspreading events where most attendees were infected (e.g. the Skagit Valley choir rehearsal).^11^ However, if human activity-dependent factors are not taken into account, H’ for all the outbreaks discussed in this paper is ∼0.1-0.5 person h^2^ m^-3^. H’ values for public indoor spaces usually fall in or near this range primarily due to similar per-person ventilation rates and public event durations. Substandard ventilation, coupled with poor air distribution, is associated with substantial increases in the risk of outbreak. However, all of the pre-pandemic example spaces analyzed are in a regime in which they are highly sensitive to mitigation efforts.

The relative risk of COVID-19 infection falls between that of two well-known airborne diseases: the more transmissible measles and the less transmissible tuberculosis. This shows that the fact that COVID-19 is less transmissible than measles does not rule out airborne transmission. These risk parameters can be applied to other airborne diseases, if outbreaks are characterized in this framework. This approach may be useful in the design and renovation of building systems. For a novel disease that was as transmissible as measles, it would be very difficult to make any indoor activities safe aside from vaccination.

Our analysis shows that mitigation measures to limit aerosol transmission are needed in most indoor spaces whenever COVID-19 is spreading in a community. Among effective measures are reducing vocalization, avoiding intense physical activities, shortening duration of occupancy, reducing number of occupants, wearing high-quality well-fitting masks, increasing ventilation, improving ventilation effectiveness and applying additional virus removal measures (such as HEPA filtration and UVGI disinfection). The use of multiple “layers of protection” is needed in many situations, while a single measure (e.g. masking) may not be able to reduce risk to low levels. We have shown that combinations of some or all of these measures are able to lower H close to 0.01 person h^2^ m^-3^, so that the expected number of secondary cases is substantially lower than 1 even in the presence of an infectious person, hence would be likely to avoid major outbreaks.

## Data Availability

All data is available in the main text or the Supporting Information.

## Acknowledgements

ZP and JLJ were partially supported by NSF AGS-1822664. TG was supported by ESRC ES/V010069/1.

## Competing interests

The authors declare no competing interests.

## Notes

### Competing Interest Statement

The authors have declared no competing interest.

### Author Declarations

This is a modeling study. Data were computationally generated.

### Summary of Updates

Significant modifications to improve the presentation of the manuscript; Monte Carlo uncertainty propagation for the attack rate vs. Hr fitting added.

