## Supporting Information for "Practical Indicators for Risk of Airborne Transmission in Shared Indoor Environments and their Application to COVID-19 Outbreaks"

### S1. Deviation of quanta concentration from steady state due to finite event duration

In case of short events where quanta concentration does not reach steady state, a correction factor, $r_{ss}$, can be introduced to account for the deviation of average quanta concentration ($c_{avg}$) from that at steady state (c):

$$r_{ss} = c_{avg} / c \qquad \text{(S1)}$$

Under the assumption of no infectious quanta in the air at the beginning of the event, $c_{avg}$ can be easily obtained from the integration of equation (1). Details of the derivation can be found elsewhere.[1,2] For a period [0, D],

$$c_{avg} = E_p f_e / (V \lambda) \times (1 - (1 - e^{-\lambda D}) / (\lambda D)) \qquad \text{(S2)}$$

Inserting equations (2) and (S2) into equations (S1) yields:

$$r_{ss} = 1 - (1 - e^{-\lambda D}) / (\lambda D) \qquad \text{(S3)}$$

The value of $r_{ss}$ as a function of λD is shown in Figure S3. $r_{ss}$ approaches to λD/2 when λD is very small and to 1 when λD is very large, and reaches 0.6 at λD ~ 2.

### S2. Monte Carlo uncertainty propagation for the fitting of attack rates vs. $H_r$

We follow the standard procedure of Monte Carlo uncertainty propagation[3] for the fitting of attack rates vs. $H_r$. We assume log-normal distributions for the variables constraining $H_r$ ($r_E$, $r_B$, D, V, and λ; $f_e$ and $f_i$ are excluded as little to no mask wearing was reported for the COVID-19 outbreaks analyzed in this study) to ensure positive values of their samples. $r_E$, $r_B$, D, V, and λ are assigned uncertainty factors of 2.5, 1.3, 1.1, 1.3, and 1.4 respectively (approximately corresponding to relative uncertainties of 150%, 30%, 10%, 30%, and 40%). The last three uncertainties are typical values for outbreak case studies. The uncertainty factor of 1.3 for $r_B$

mainly reflects the possible error arising from the discretization of physical intensity levels in the [4] dataset. We assume an uncertainty factor of 2.5 for $r_E$ because Buonanno et al.[5] estimated the uncertainty of $E_{p0}xr_E$ for COVID-19 to be an order of magnitude and we think that $r_E$, a relative factor that depends largely on type of activity but not on that of disease, contributes only a minority of this uncertainty. Since attack rate (AR) is bounded between 0 and 100%, it does not follow a log-normal distribution. We use a similar transformation as in Gans et al.[6], i.e., AR / (1 - AR), to expand the domain of the samples from [0, 1] to [0, +∞). The intermediate samples then can be depicted with a log-normal distribution. We assign an uncertainty factor of 1.1 to the intermediate samples. The generated samples are then reversely transformed into the AR samples. When AR is small, the assigned uncertainty factor of 1.1 approximately corresponds to a relative uncertainty of 10% for AR; while when AR is close to 1, this uncertainty factor reflects an approximate relative uncertainty of 10% for non-attack rate, i.e., (1 - AR). 10000 random samples of $r_E$, $r_B$, D, V, λ, and AR are generated for each of the COVID-19 case studies in Table 1. A fitting can be done for one sample of $r_E$, $r_B$, D, V, λ, and AR of all those case studies, yielding a sample of the fitted parameter, $E_{p0}$. This fitting is repeated for all 10000 samples of the input parameters, giving 10000 samples of $E_{p0}$, apparently log-normally distributed, with 5$^{th}$ and 95$^{th}$ percentiles being 8.7 and 45.4 quanta $h^{-1}$, respectively.

**Tables**

| *Symbol* | *Physical meaning* | *Unit (dimension -less if no unit indicated)* |
|---|---|---|
| B | Volumetric breathing rate of a susceptible person | $m^3 h^{-1}$ |
| $B_0$ | Volumetric breathing rate of a resting susceptible person | $m^3 h^{-1}$ |
| c | Virus concentration | quanta $m^{-3}$ |
| $c_{avg}$ | Average virus concentration in the air over the duration of the event | quanta $m^{-3}$ |
| D | Duration of the event | h |
| $E_p$ | SARS-CoV-2 exhalation rate by an infector | quanta $h^{-1}$ |
| $E_{p0}$ | SARS-CoV-2 exhalation rate by an infector resting and only orally breathing | quanta $h^{-1}$ |
| $f_e$ | Exhalation penetration efficiency for face covering | |
| $f_i$ | Inhalation penetration efficiency for face covering | |
| H | Infection risk parameter, as defined in equation (11) | persons $h^2 m^{-3}$ |
| H’ | Infection risk parameter without activity taken into account, as defined in equation (14) | persons $h^2 m^{-3}$ |

| | | |
|---|---|---|
| $H_r$ | Relative infection risk parameter, as defined in equation (15) | $h^2\ m^{-3}$ |
| $\eta_I$ | Probability of an occupant being an infector | |
| $\lambda$ | First-order overall rate constant of the virus infectivity loss | $h^{-1}$ |
| $\lambda_0$ | Ventilation rate | $h^{-1}$ |
| $\lambda_{cle}$ | Virus removal rate by cleaning devices | $h^{-1}$ |
| $\lambda_{dec}$ | Virus infectivity decay rate | $h^{-1}$ |
| $\lambda_{dep}$ | Deposition rate of airborne virus-containing particles onto surfaces | $h^{-1}$ |
| L | Ventilation rate per susceptible person | liter $s^{-1}$ person$^{-1}$ |
| N | Number of occupants | |
| $N_i$ | Number of infectors | |
| $N_{sus}$ | Number of susceptible persons | |
| $N_{si}$ | Number of secondary infections | |
| n | Amount of the virus infectious doses inhaled by a susceptible person in a given indoor environment | quanta |
| P | Probability of infection of a susceptible person conditional on the presence of an infector | |
| $P_a$ | Absolute probability of infection of a susceptible person | |

| | | |
|---|---|---|
| $r_{ss}$ | Ratio of the average virus concentration to that at steady state | |
| $r_B$ | Relative breathing rate enhancement factor (vs. $B_0$) for an activity | |
| $r_E$ | Relative virus exhalation rate enhancement factor (vs. $E_{p0}$) for an activity | |
| V | Indoor environment volume | $m^3$ |

*Table S1: mathematical symbols used in this study*

90 (a)

| Activity | | Relative quanta emission rate factor |
|---|---|---|
| Physical intensity | Vocalization | |
| Resting | Oral breathing | 1 |
| | Speaking | 4.7 |
| | Loudly speaking | 30.3 |
| Standing | Oral breathing | 1.2 |
| | Speaking | 5.7 |
| | Loudly speaking | 32.6 |
| Light exercise | Oral breathing | 2.8 |
| | Speaking | 13.2 |
| | Loudly speaking | 85 |
| Moderate exercise | Oral breathing | 4.3 |
| | Speaking | 20.4 |
| | Loudly speaking | 132 |
| Heavy exercise | Oral breathing | 6.8 |
| | Speaking | 31.6 |
| | Loudly speaking | 204 |

(b)

| Age group (year) | Activity level | | | | |
|---|---|---|---|---|---|
| | Sleep or nap | Sedentary /passive | Light intensity | Moderate intensity | High intensity |
| <1 | 0.63 | 0.64 | 1.6 | 2.9 | 5.4 |
| 1 - <2 | 0.94 | 1.0 | 2.5 | 4.4 | 7.9 |
| 2 - <3 | 0.96 | 1.0 | 2.5 | 4.4 | 8.1 |
| 3 - <6 | 0.90 | 0.94 | 2.3 | 4.4 | 7.7 |
| 6 - <11 | 0.94 | 1.0 | 2.3 | 4.6 | 8.7 |
| 11 - <16 | 1.0 | 1.1 | 2.7 | 5.2 | 10 |
| 16 - <21 | 1.0 | 1.1 | 2.5 | 5.4 | 10 |
| 21 - <31 | 0.90 | 0.88 | 2.5 | 5.4 | 10 |
| 31 - <41 | 1.0 | 0.89 | 2.5 | 5.6 | 10 |
| 41 - <51 | 1.0 | 1.0 | 2.7 | 5.8 | 11 |
| 51 - <61 | 1.1 | 1.0 | 2.7 | 6.0 | 11 |
| 61 - <71 | 1.1 | 1.0 | 2.5 | 5.4 | 9.8 |
| 71 - <81 | 1.1 | 1.0 | 2.5 | 5.2 | 9.8 |

| | | | | | |
|---|---|---|---|---|---|
| ≥81 | 1.1 | 1.0 | 2.5 | 5.2 | 10 |
| Average | 1.0 | 1.0 | 2.4 | 5.0 | 9 |

*Table S2: relative factors of (a) quanta emission and (b) volumetric breathing rates for different activities according to refs [5,7] and ref [4], respectively. The values of relative quanta emission rate factor for moderate exercise in (a) are interpolated as in ref [2].*

| | | |
|---|---|---|
| **Relative quanta emission factor** | | |
| Silent | 1 | |
| Speaking | 5 | |
| Shouting, singing | 30 | |
| Heavy exercise | 7 | |
| **Relative breathing rate factor** | | |
| Silent | 1 | |
| Speaking | 1 | |
| Shouting, singing | 1 | |
| Heavy exercise | 10 | |
| **Low occupancy** | 10 | persons |
| **High occupancy** | 100 | persons |
| **Ventilation rate** | | |
| Outdoor and well ventilated | 500 | ACH |
| Indoor and well ventilated | 10 | ACH |
| Poorly ventilated | 1 | ACH |
| **Face coverings** | | |
| Exhalation filtration efficiency | 50% | |
| Inhalation filtration efficiency | 30% | |
| **Contact time** | | |
| Short | 1 | h |
| Long | 10 | h |
| **Effective volume** | | |
| Indoor | 300 | $m^3$ |
| Outdoor | 300 | $m^3$ |

*Footnote: A rough estimate can be obtained as follows. To be comparable with the indoor volume, the outdoor volume is assumed to be the same as the indoor one (10 m x 10 m x 3 m box). The outdoor ventilation rate corresponds to the ventilation by wind passing through a horizontal dimension of the outdoor box (10 m) at 5 km $h^{-1}$ (~1.4 m $s^{-1}$, toward the low end of the monthly mean wind speed in US cities).[8] The outdoor box dimensions and wind* *speed are input parameters for the table in the same format in the COVID-19 Aerosol Transmission Estimator (Figure S2) for its users to more easily estimate equivalent outdoor ventilation.*

*Table S3: values of the parameters used for computation of Table 2 in the main paper.*

| Indoor environment type | $r_E$ | $r_B$ | $f_e x f_i$ | D (h) | $N_{sus}$ | V ($m^3$) | $\lambda_0 + \lambda_{cle}$ ($h^{-1}$) | $r_{ss}$ | H (persons $h^2$ $m^{-3}$) | $H_r$ ($h^2$ $m^{-3}$) | Predicted number of secondary cases |
|---|---|---|---|---|---|---|---|---|---|---|---|
| ASHRAE standard cases | | | | | | | | | | | |
| Prison dayroom | 2.8 | 2.4 | 1 | 8 | 300 | 5.0E+03 | 0.76 | 0.84 | 3.6E+00 | 1.2E-02 | 1.8E+01 |
| | 2.8 | 2.4 | 0.35 | 4 | 150 | 5.0E+03 | 3.8 | 0.93 | 7.0E-02 | 4.7E-04 | 3.8E-01 |
| Middle school classroom | 1 | 1.1[a] | 1 | 5 | 20 | 1.7E+02 | 2.8 | 0.93 | 2.1E-01 | 1.1E-02 | 1.1E+00 |
| | 1 | 1.1[a] | 0.35 | 2.5 | 10 | 1.7E+02 | 5.8 | 0.93 | 9.0E-03 | 9.0E-04 | 4.8E-02 |
| Concert hall/theater | 85 | 2.4 | 1 | 2 | 300 | 1.3E+04 | 0.49 | 0.36 | 7.0E+00 | 2.3E-02 | 3.5E+01 |
| | 85 | 2.4 | 0.35 | 1 | 150 | 1.3E+04 | 3.5 | 0.72 | 1.7E-01 | 1.1E-03 | 9.2E-01 |
| Restaurant | 4.7 | 1 | 1 | 1 | 50 | 2.1E+02 | 4.3 | 0.77 | 2.0E-01 | 3.9E-03 | 1.0E+00 |
| | 2.9[b] | 1 | 1[c] | 1[d] | 25 | 2.1E+02 | 7.3 | 0.86 | 4.0E-02 | 1.6E-03 | 2.1E-01 |
| Hotel lobbies/prefunction | 2.8 | 2.4 | 1 | 8 | 50 | 1.0E+03 | 0.86 | 0.86 | 2.7E+00 | 5.3E-02 | 1.2E+01 |
| | 2.8 | 2.4 | 0.35 | 4 | 25 | 1.0E+03 | 3.9 | 0.94 | 5.7E-02 | 2.3E-03 | 3.0E-01 |
| Airport terminal /railway station | 2.8 | 2.4 | 1 | 1 | 1000 | 1.0E+04 | 1.5 | 0.48 | 2.2E-01 | 2.2E-04 | 1.2E+00 |
| | 2.8 | 2.4 | 0.35 | 1[d] | 500 | 1.0E+04 | 4.5 | 0.78 | 2.0E-02 | 4.1E-05 | 1.1E-01 |
| Hospital general | 50[e] | 1 | 0.35[f] | 8 | 20 | 3.0E+02 | 1.6 | 0.92 | 5.3E+00 | 2.6E-01 | 1.5E+01 |

| | | | | | | | | | | | |
|---|---|---|---|---|---|---|---|---|---|---|---|
| examination room | 50[e] | 1 | 0.01[g] | 4 | 10 | 3.0E+02 | 9.0[h] | 0.97 | 7.2E-03 | 7.2E-04 | 3.9E-02 |
| Library | 1 | 1 | 1 | 2 | 100 | 3.0E+03 | 1.0 | 0.57 | 3.7E-02 | 3.7E-04 | 2.0E-01 |
| | 1 | 1 | 0.35 | 2[d] | 50 | 3.0E+03 | 4.0 | 0.88 | 2.5E-03 | 5.1E-05 | 1.4E-02 |
| Museum/gallery | 1.2 | 1.5 | 1 | 2 | 200 | 5.0E+03 | 0.66 | 0.44 | 9.7E-02 | 4.9E-04 | 5.2E-01 |
| | 1.2 | 1.5 | 0.35 | 2[d] | 100 | 5.0E+03 | 3.7 | 0.86 | 6.0E-03 | 6.0E-05 | 3.2E-02 |
| Place of religious worship | 30 | 1 | 1 | 2 | 100 | 8.3E+02 | 1.2 | 0.62 | 3.8E+00 | 3.8E-02 | 1.8E+01 |
| | 4.7[i] | 1 | 0.35 | 1 | 50 | 8.3E+02 | 4.2 | 0.76 | 1.8E-02 | 3.6E-04 | 9.6E-02 |
| Mall common area | 2.8 | 2.4 | 1 | 2 | 500 | 7.5E+03 | 1.1 | 0.59 | 4.9E-01 | 9.7E-04 | 2.6E+00 |
| | 2.8 | 2.4 | 0.35 | 1 | 250 | 7.5E+03 | 4.1 | 0.76 | 1.5E-02 | 5.8E-05 | 7.8E-02 |
| Supermarket | 2.8 | 2.4 | 1 | 8 | 100 | 7.5E+03 | 0.36 | 0.67 | 1.3E+00 | 1.3E-02 | 6.9E+00 |
| | 2.8 | 2.4 | 0.35 | 4 | 50 | 7.5E+03 | 3.4 | 0.93 | 1.7E-02 | 3.5E-04 | 9.2E-02 |
| Gym, sports arena (play area) | 6.8 | 9 | 1 | 1 | 100 | 1.4E+04 | 0.58 | 0.24 | 1.8E-01 | 1.8E-03 | 9.5E-01 |
| | 6.8 | 9 | 0.35 | 0.5 | 50 | 1.4E+04 | 3.6 | 0.53 | 5.6E-03 | 1.1E-04 | 3.0E-02 |
| Other cases | | | | | | | | | | | |
| Physical education class | 6.8 | 9 | 1 | 1 | 30 | 1.3E+03 | 1.9 | 0.56 | 4.1E-01 | 1.4E-02 | 2.1E+00 |
| | 6.8 | 9 | 0.35 | 0.5 | 15 | 1.4E+04[j] | 4.9 | 0.63 | 1.4E-03 | 9.6E-05 | 7.7E-03 |
| Subway car[k] | 1 | 1 | 1 | 0.33 | 30 | 1.5E+02 | 5.7 | 0.55 | 6.5E-03 | 2.2E-04 | 3.5E-02 |

| | | | | | | | | | | | |
|---|---|---|---|---|---|---|---|---|---|---|---|
| | 1 | 1 | 0.35 | 0.33[d] | 30[l] | 1.5E+02 | 9.3[m] | 0.69 | 1.7E-03 | 5.8E-05 | 9.3E-03 |
| Large family dinner | 4.7 | 1 | 1 | 2 | 12 | 3.0E+02 | 0.5 | 0.37 | 2.8E-01 | 2.3E-02 | 1.4E+00 |
| | 2.9[b] | 1 | 0.35 | 2[d] | 6 | 3.0E+02 | 3.5 | 0.86 | 9.9E-03 | 1.7E-03 | 5.3E-02 |
| Shared office | 4.7 | 1 | 1 | 8 | 2 | 3.4E+01 | 2 | 0.94 | 1.0E+00 | 5.2E-01 | 1.9E+00 |
| | 1.7[n] | 1 | 0.35 | 4 | 1 | 3.4E+01 | 5 | 0.95 | 1.3E-02 | 1.3E-02 | 6.9E-02 |
| Large university classroom[o] | 30 | 1 | 1 | 1 | 150 | 7.0E+02 | 2 | 0.57 | 1.8E+00 | 1.2E-02 | 9.5E+00 |
| | 4.7[i] | 1 | 0.35 | 1[c] | 60[p] | 7.0E+02 | 11[q] | 0.91 | 1.2E-02 | 1.9E-04 | 6.2E-02 |
| University laboratory | 2.8 | 2.4 | 1 | 8 | 10 | 2.3E+02 | 6 | 0.98 | 3.9E-01 | 3.9E-02 | 1.9E+00 |
| | 2.8 | 2.4 | 0.35 | 4 | 3[o] | 2.3E+02 | 9 | 0.97 | 1.3E-02 | 4.5E-03 | 7.1E-02 |
| Outbreaks | | | | | | | | | | | |
| Guangzhou restaurant | 9.3[r] | 1 | 1 | 1.2 | 20 | 9.7E+01 | 0.67 | 0.31 | 1.1E+00 | 5.4E-02 | 5.0E+00 |
| | 4.7[s] | 1 | 1[c] | 0.6 | 10 | 9.7E+01 | 3.67 | 0.60 | 4.7E-02 | 4.7E-03 | 2.5E-01 |
| Skagit Choir | 85 | 2.5[t] | 1 | 2.5 | 60 | 8.1E+02 | 0.7 | 0.53 | 3.0E+01 | 5.0E-01 | 5.6E+01 |
| | 85 | 2.5[t] | 0.35 | 1.3 | 30 | 8.1E+02 | 3.7 | 0.79 | 7.3E-01 | 2.4E-02 | 3.7E+00 |

*Footnotes: [a] for sedentary teenagers; [b] half resting - oral breathing + half resting - speaking; [c] no face covering; [d] no duration reduction; [e] for a coughing infector (see Footnote e of Table 1 for detail of the estimation); [f] use of surgical masks for the pre-pandemic setting; [g] N95 respirators and fit tests required (resulting in $f_e$ and $f_i$ of 0.1) before allowed indoors; [h] ventilation rate increased to 6 $h^{-1}$; [i] reduction of vocalization level from loudly speaking to speaking (with the aid of, for example, microphone); [j] use of a much larger room if the event has to be indoors; [k] real-world case;[2] [l] no occupancy reduction; [m] $\lambda_{cle}$ = 3.6 $h^{-1}$; [n] 4/5 resting - oral breathing + 1/5 resting - speaking; [o] real-world case;[9] [p] occupancy reduction larger than 50%; [q] ventilation rate increased to the maximum and no additional virus removal* *applied; [r] talking during half of the time and half normal / half loud talking assumed; [s] resting - speaking; [t] light intensity for 61-<71 years.*

*Table S4: parameters for pre-pandemic use of various indoor spaces, and for possible lower-risk scenarios while COVID-19 is active (rows in gray). The predicted number of secondary cases is estimated based on the fitted trend in Figure 1b. The ventilation rates ($\lambda_0$) of the ASHRAE standard cases correspond to the minimum requirement recommended in ref [10]. The other cases are based on real-world indoor spaces or reasonable estimation. $r_E$ and $r_B$ are estimated mostly based on the typical values for all-age-group averages in Table S2. No additional virus-removal devices or no face covering are used in the pre-pandemic cases. Common measures for the lower-risk scenarios in this table are half occupancy, half duration, surgical mask wearing ($f_e \times f_i = 0.35$),[2,11] and use of additional virus-removing devices (e.g. HEPA filter) with $\lambda_{cle} = 3\ h^{-1}$. Two literature outbreaks in Table 1, i.e. the Guangzhou restaurant[12] and Skagit Choir[1] cases, are also shown for comparison. See footnotes for the exceptions to these descriptions.*

### Figures

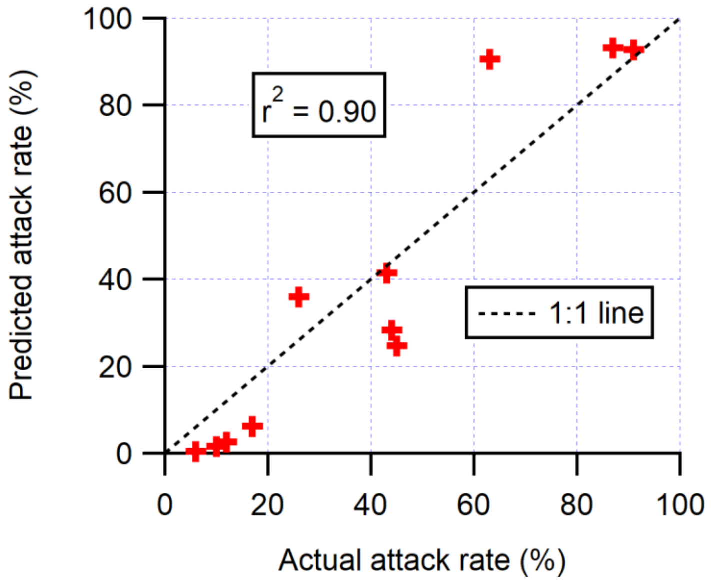

*Figure S1: Attack rates of the COVID-19 outbreaks shown in Table 1 predicted according to the fitted trend line in Figure 2b vs. actual attack rates of those outbreaks. The correlation coefficient between the two types of attack rates and the 1:1 line are also shown.*

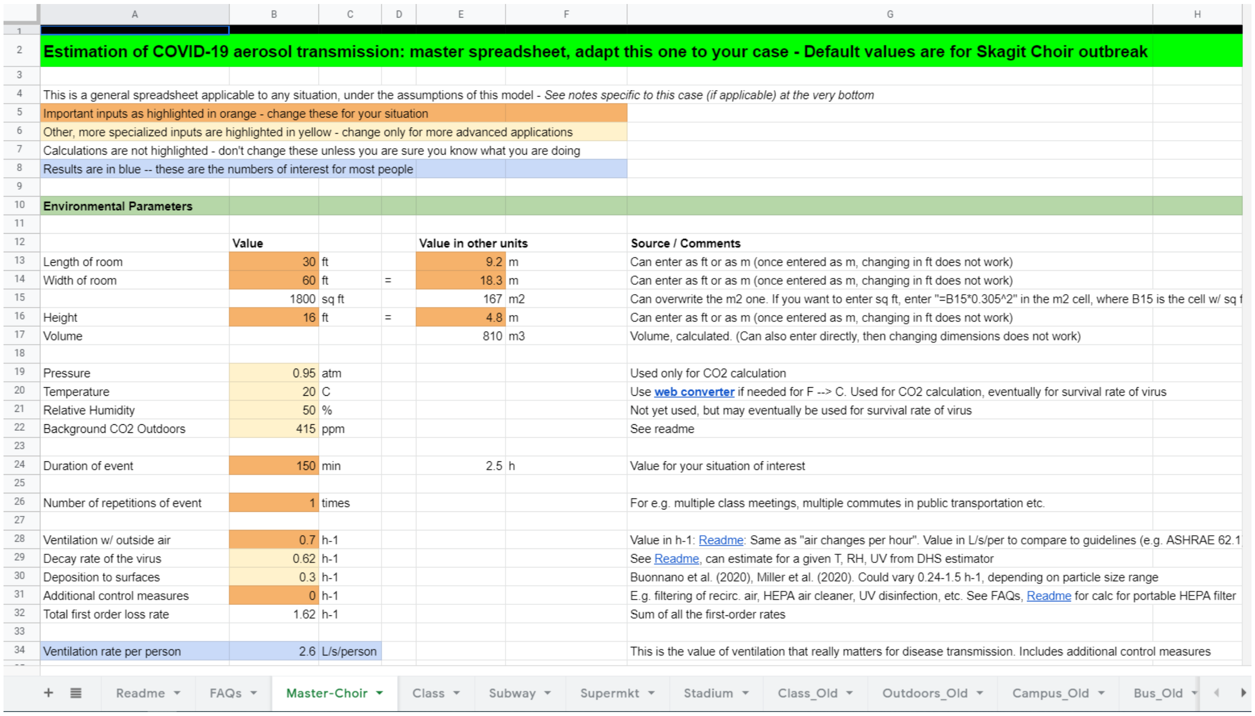

| | A | B | C | D | E | F | G |
|---|---|---|---|---|---|---|---|
| 2 | **Estimation of COVID-19 aerosol transmission: master spreadsheet, adapt this one to your case - Default values are for Skagit Choir outbreak** | | | | | | |
| 4 | This is a general spreadsheet applicable to any situation, under the assumptions of this model - *See notes specific to this case (if applicable) at the very bottom* | | | | | | |
| 5 | Important inputs as highlighted in orange - change these for your situation | | | | | | |
| 6 | Other, more specialized inputs are highlighted in yellow - change only for more advanced applications | | | | | | |
| 7 | Calculations are not highlighted - don't change these unless you are sure you know what you are doing | | | | | | |
| 8 | Results are in blue -- these are the numbers of interest for most people | | | | | | |
| 10 | **Environmental Parameters** | | | | | | |
| 12 | | **Value** | | | **Value in other units** | | **Source / Comments** |
| 13 | Length of room | 30 | ft | | 9.2 | m | Can enter as ft or as m (once entered as m, changing in ft does not work) |
| 14 | Width of room | 60 | ft | = | 18.3 | m | Can enter as ft or as m (once entered as m, changing in ft does not work) |
| 15 | | 1800 | sq ft | | 167 | m2 | Can overwrite the m2 one. If you want to enter sq ft, enter "=B15*0.305^2" in the m2 cell, where B15 is the cell w/ sq f |
| 16 | Height | 16 | ft | = | 4.8 | m | Can enter as ft or as m (once entered as m, changing in ft does not work) |
| 17 | Volume | | | | 810 | m3 | Volume, calculated. (Can also enter directly, then changing dimensions does not work) |
| 19 | Pressure | 0.95 | atm | | | | Used only for CO2 calculation |
| 20 | Temperature | 20 | C | | | | Use **web converter** if needed for F --> C. Used for CO2 calculation, eventually for survival rate of virus |
| 21 | Relative Humidity | 50 | % | | | | Not yet used, but may eventually be used for survival rate of virus |
| 22 | Background CO2 Outdoors | 415 | ppm | | | | See readme |
| 24 | Duration of event | 150 | min | | 2.5 | h | Value for your situation of interest |
| 26 | Number of repetitions of event | 1 | times | | | | For e.g. multiple class meetings, multiple commutes in public transportation etc. |
| 28 | Ventilation w/ outside air | 0.7 | h-1 | | | | Value in h-1: Readme: Same as "air changes per hour". Value in L/s/per to compare to guidelines (e.g. ASHRAE 62.1 |
| 29 | Decay rate of the virus | 0.62 | h-1 | | | | See Readme, can estimate for a given T, RH, UV from DHS estimator |
| 30 | Deposition to surfaces | 0.3 | h-1 | | | | Buonnano et al. (2020), Miller et al. (2020). Could vary 0.24-1.5 h-1, depending on particle size range |
| 31 | Additional control measures | 0 | h-1 | | | | E.g. filtering of recirc. air, HEPA air cleaner, UV disinfection, etc. See FAQs, Readme for calc for portable HEPA filter |
| 32 | Total first order loss rate | 1.62 | h-1 | | | | Sum of all the first-order rates |
| 34 | Ventilation rate per person | 2.6 | L/s/person | | | | This is the value of ventilation that really matters for disease transmission. Includes additional control measures |

*Figure S2: screenshot of the COVID-19 Aerosol Transmission Estimator.[13] The top of the sheet simulating the Skagit Valley choir outbreak is shown.*

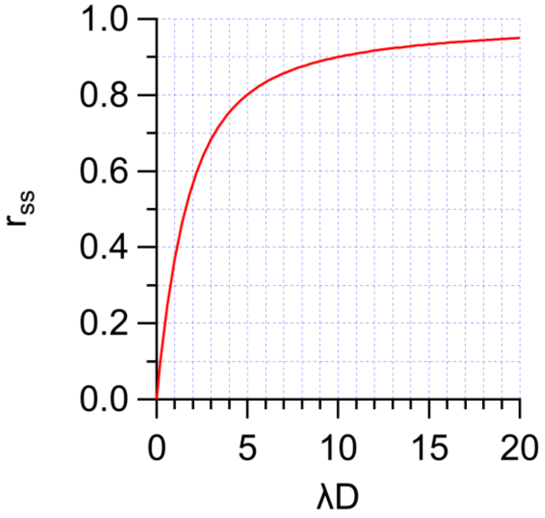

*Figure S3: Ratio of the quanta concentration averaged over a period [0, D] to that at steady state ($r_{ss}$) as a function of the product of total first-order quanta loss rate constant ($\lambda$) and the event duration (D).*
